# Immune Cell Profiling of COVID-19 Patients in the Recovery Stage by Single-Cell Sequencing

**DOI:** 10.1101/2020.03.23.20039362

**Authors:** Wen Wen, Wenru Su, Hao Tang, Wenqing Le, Xiaopeng Zhang, Yingfeng Zheng, Xiuxing Liu, Lihui Xie, Jianmin Li, Jinguo Ye, Xiuliang Cui, Yushan Miao, Depeng Wang, Jiantao Dong, Chuanle Xiao, Wei Chen, Hongyang Wang

## Abstract

COVID-19, caused by SARS-CoV-2, has recently affected over 300,000 people and killed more than 10,000. The manner in which the key immune cell subsets change and their states during the course of COVID-19 remain unclear. Here, we applied single-cell technology to comprehensively characterize transcriptional changes in peripheral blood mononuclear cells during the recovery stage of COVID-19. Compared with healthy controls, in patients in the early recovery stage (ERS) of COVID-19, T cells decreased remarkably, whereas monocytes increased. A detailed analysis of the monocytes revealed that there was an increased ratio of classical CD14^++^ monocytes with high inflammatory gene expression as well as a greater abundance of CD14^++^IL1B^+^ monocytes in the ERS. CD4^+^ and CD8^+^ T cells decreased significantly and expressed high levels of inflammatory genes in the ERS. Among the B cells, the plasma cells increased remarkably, whereas the naïve B cells decreased. Our study identified several novel B cell-receptor (BCR) changes, such as IGHV3-23 and IGHV3-7, and confirmed isotypes (IGHV3-15, IGHV3-30, and IGKV3-11) previously used for virus vaccine development. The strongest pairing frequencies, IGHV3-23-IGHJ4, indicated a monoclonal state associated with SARS-CoV-2 specificity. Furthermore, integrated analysis predicted that IL-1β and M-CSF may be novel candidate target genes for inflammatory storm and that TNFSF13, IL-18, IL-2 and IL-4 may be beneficial for the recovery of COVID-19 patients. Our study provides the first evidence of an inflammatory immune signature in the ERS, suggesting that COVID-19 patients are still vulnerable after hospital discharge. Our identification of novel BCR signaling may lead to the development of vaccines and antibodies for the treatment of COVID-19.

**Highlights:**

- The immune response was sustained for more than 7 days in the early recovery stage of COVID-19, suggesting that COVID-19 patients are still vulnerable after hospital discharge.
- Single-cell analysis revealed a predominant subset of CD14^++^ IL1β^+^ monocytes in patients in the ERS of COVID-19.
- Newly identified virus-specific B cell-receptor changes, such as IGHV3-23, IGHV3-7, IGHV3-15, IGHV3-30, and IGKV3-11, could be helpful in the development of vaccines and antibodies against SARS-CoV-2.
- IL-1β and M-CSF were discovered as novel mediators of inflammatory cytokine storm, and TNFSF13, IL-2, IL-4, and IL-18 may be beneficial for recovery.

## Introduction

COVID-19, caused by severe acute respiratory syndrome coronavirus 2 (SARS-CoV-2), has spread in many countries ^[1-3]^. As of March 21, 2020, SARS-CoV-2 has affected over 300,000 people and killed more than 10,000 of those affected in more than 160 countries. Following its global spread, the World Health Organization declared it a public health emergency of international concern ^[4]^. COVID-19 shows symptoms of fever, dry cough, fatigue, diarrhea, conjunctivitis, and pneumonia. Some patients develop severe pneumonia, acute respiratory distress syndrome (ARDS), or multiple organ failure ^[5-7]^. Although scientists and clinicians worldwide have made great efforts to produce vaccines and explored antiviral drugs ^[8, 9]^, there is still no specific medicine and effective clinical treatment for COVID-19 ^[10,11]^.

Immune system dysregulation, such as lymphopenia and inflammatory cytokine storm, have been observed and are believed to be associated with the severity of pathogenic coronavirus infections, such as severe acute respiratory syndrome coronavirus (SARS-CoV) and Middle East respiratory syndrome coronavirus (MERS-CoV) infections ^[12, 13]^. With regard to COVID-19, recent studies also found decreases in lymphocyte numbers and increases in serum inflammatory cytokine levels in peripheral blood ^[5, 14]^. However, the manner in which key immune cell subsets change and their states during COVID-19 have remained largely unclear. Thus, defining these key cellular subsets and their states in COVID-19 is a crucial step in obtaining critical insights into the immune clearance mechanism and developing new therapeutic strategies for COVID-19.

Here, we applied single-cell RNA sequencing (scRNA-seq) to comprehensively characterize the changes in peripheral blood mononuclear cells (PBMCs) from 10 COVID-19 patients. Our study depicts a high-resolution transcriptome landscape of blood immune cell subsets during the recovery stage of COVID-19. It reveals that, compared to that in the healthy controls (HCs), monocytes containing high inflammatory gene expression and IL1β^+^ subsets predominated, whereas CD4^+^ T cells decreased remarkably in patients in the early recovery stage of COVID-19. We found that T and B cell clones were highly expanded during the recovery stage in COVID-19 patients. Furthermore, several specific BCR changes in COVID-19 patients during the recovery stage may be helpful for vaccine and antibody production.

## Results

### Study design and analysis of single immune cell profiling in COVID-19 patients

To map the immune microenvironment of COVID-19 patients, we identified mirroring changes in the blood and pinpointed cell-specific alterations associated with disease severity and recovery; we then integrated single-cell RNA sequencing (scRNA-seq), single-cell paired BCR, and single-cell paired TCR analysis from a total of 10 COVID-19 patients in the early recovery stage (ERS) or late recovery stage (LRS) (70,858 PBMCs). We also collected scRNA-seq data (57,238 cells) from five healthy donors as controls (Fig. 1A and Fig. S1). This dataset passed stringent high-quality filtering. Single-cell suspensions of the scRNA-seq samples were converted to barcoded scRNA-seq libraries using 10X Genomics. CellRanger software (version 3.1.0) was used for the initial processing of the sequencing data.

**Figure 1.**
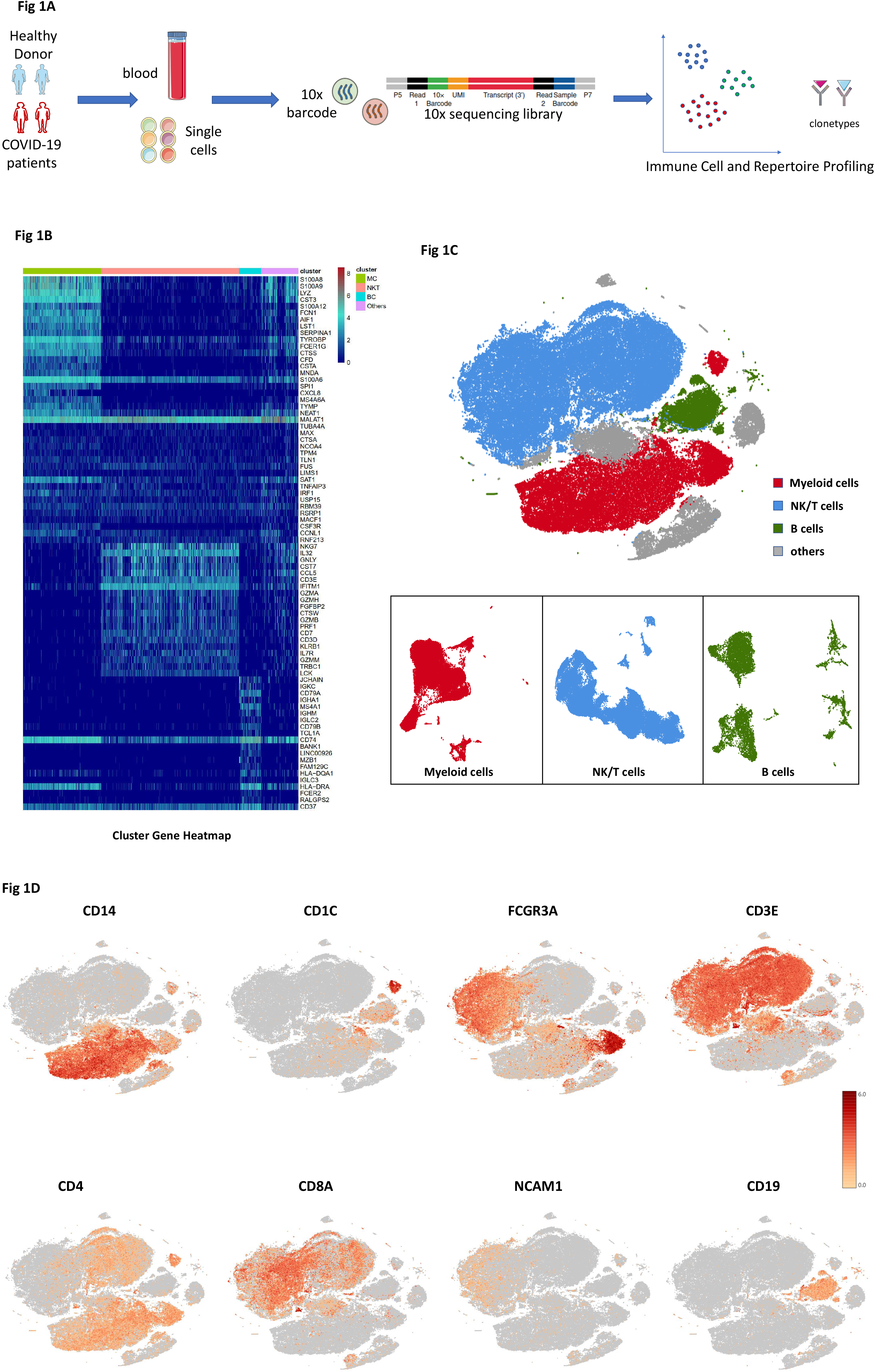
Study design and analysis of single immune cell profiling in COVID-19 patients. A. Schematics of the experimental design for single-cell RNA (sc-RNA) sequencing. Peripheral blood mononuclear cells (PBMCs) were collected from COVID-19 patients and healthy controls (HCs) and then processed via sc-RNA, sc-BCR, and sc-TCR sequencing using the 10X-Based Genomics platform. B. The heatmaps show differentially expressed genes (DEGs) upregulated in myeloid cells, NKT cells, B cells, and other clusters of PBMCs. C. t-distributed stochastic neighbor embedding (t-SNE) plot showing myeloid cells (red), NKT cells (blue), B cells (green), and other clusters (gray) of PBMCs identified using integrated and classification analysis. D. t-SNE projection of canonical markers, including *CD14, CD1C*, and *FCGR3A* for myeloid cells; *CD3E, CD4, CD8A*, and *NCAM1* for NKT cells; and *CD19* for B cells as indicated in the legend.

Using t-distributed stochastic neighbor embedding (t-SNE), we analyzed the distribution of the three immune cell lineages, myeloid, NKT, and B cells, based on the expression of canonical lineage markers and other genes specifically upregulated in each cluster (Fig. 1B, C). For marker genes, expression values in each cell positioned in a t-SNE are shown in Fig. 1D. We next clustered the cells of each lineage separately and identified a total of 20 immune cell clusters.

### An overview of NKT, B, and myeloid cells in the blood of convalescent patients with COVID-19

The immune cell compartment of patients who have recovered from COVID-19 infection comprised all major immune lineages. We analyzed 128,096 scRNA-seq profiles that passed quality control, including 36,442 myeloid cells, 64,247 NKT cells, and 10,177 B cells from five HCs, five ERS, and five LRS patients. The sketchy clustering analysis landscape of each subject is presented in Fig. S2A, and the merged image of each group is shown in Fig. 2A. We discovered that COVID-19 patients, including ERS and LRS, demonstrated a higher proportion of myeloid cells compared to the HCs, but with a lower proportion of NKT cells (Fig. 2B, C). Interestingly, LRS patients had more B cells and NKT cells, but less myeloid cells, than the ERS patients (Fig. 2B, C). Thus, these findings indicated that COVID-19 patients had decreased lymphocyte counts and increased counts of myeloid cells in peripheral blood.

**Figure 2.**
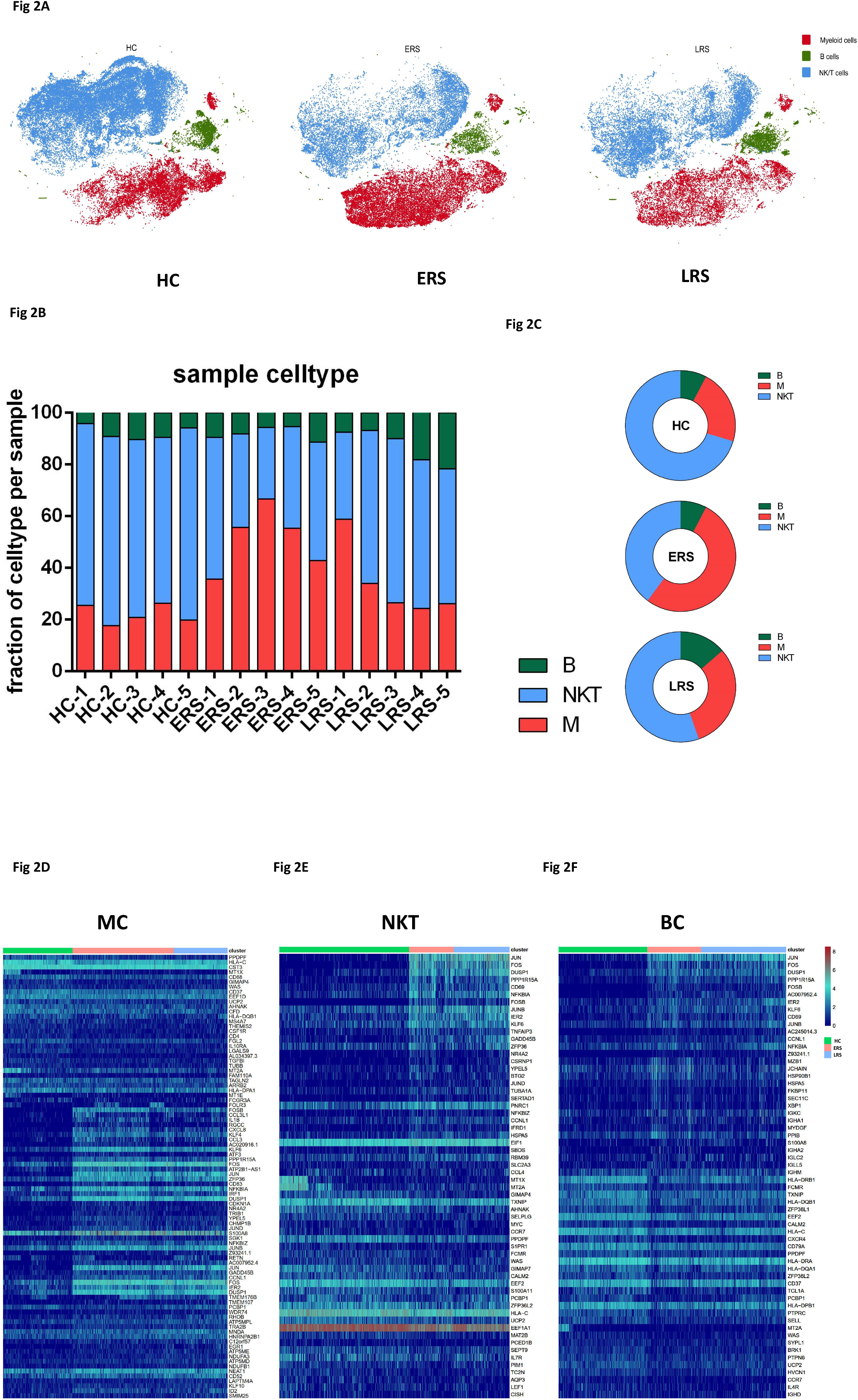
An overview of NKT, B, and myeloid cells in the blood of convalescent patients with COVID-19. A. The t-SNE plot shows a comparison of the clustering distribution across HCs as well as early recovery stage (ERS) and late recovery stage (LRS) patients with COVID-19. B. The bar plot shows the relative contributions of myeloid, NKT, and B cells by individual samples, including five HCs, five ERS patients, and five LRS patients. C. The pie chart shows the percentages of myeloid, NKT, and B cells across HCs as well as ERS and LRS patients with COVID-19. D. The heatmaps show the DEGs of myeloid cells among the HCs and the ERS and LRS COVID-19 patients. E. The heatmaps show the DEGs of NKT cells among the HCs and the ERS and LRS COVID-19 patients. F. The heatmaps show the DEGs of B cells among the HCs and the ERS and LRS COVID-19 patients.

To further understand the changes in the myeloid, NKT, and B cells in COVID-19 patients, we conducted differential expression gene (DEG) analysis of the NKT, B, and myeloid cells between the HCs and patients. The heat maps are shown in Fig. 2D-F. Inflammatory cytokines and chemokines such as *IL1B, CCL3, IRF1, DUSP1, JUN*, and *FOS* were all expressed at high levels in patients, regardless of myeloid cells (Fig. 2D), NKT cells (Fig. 2E), or B cells (Fig. 2F).

Collectively, our results demonstrated that myeloid cells increased, whereas NKT cells decreased in the peripheral blood of COVID-19 patients and that the immune cell compositions differed between the patients in the ERS and LRS.

### Myeloid cell subsets and their states in the blood of convalescent patients with COVID-19

To further understand the changes in the monocytes in patients in the early and late recovery stages of COVID-19, we conducted gene expression analysis and sub-clustered the myeloid cells into six transcriptionally distinct subsets using Uniform Manifold Approximation and Projection (UMAP). Classical CD14^++^ monocytes (M1), non-classical CD16^++^ (FCGR3A) CD14^-/+^ monocytes (M2), intermediate CD14^++^ CD16^+^ monocytes (M3), CD1C^+^ cDC2 (M4), CLEC9A^+^ cDC1 (M5), and pDC (CLEC4C^+^CD123^+^) (M6) were present in the six distinct clusters (Fig. 3A, B, and S). We found that the compartment of the monocyte subset differed remarkably among the HCs and COVID-19 patients (Fig. 3C). Among the myeloid cells, the ratio of classical CD14^++^ monocytes (M1) higher in the ERS patients than in the HCs and was almost normal in the LRS patients (Fig. 3C).

**Figure 3.**
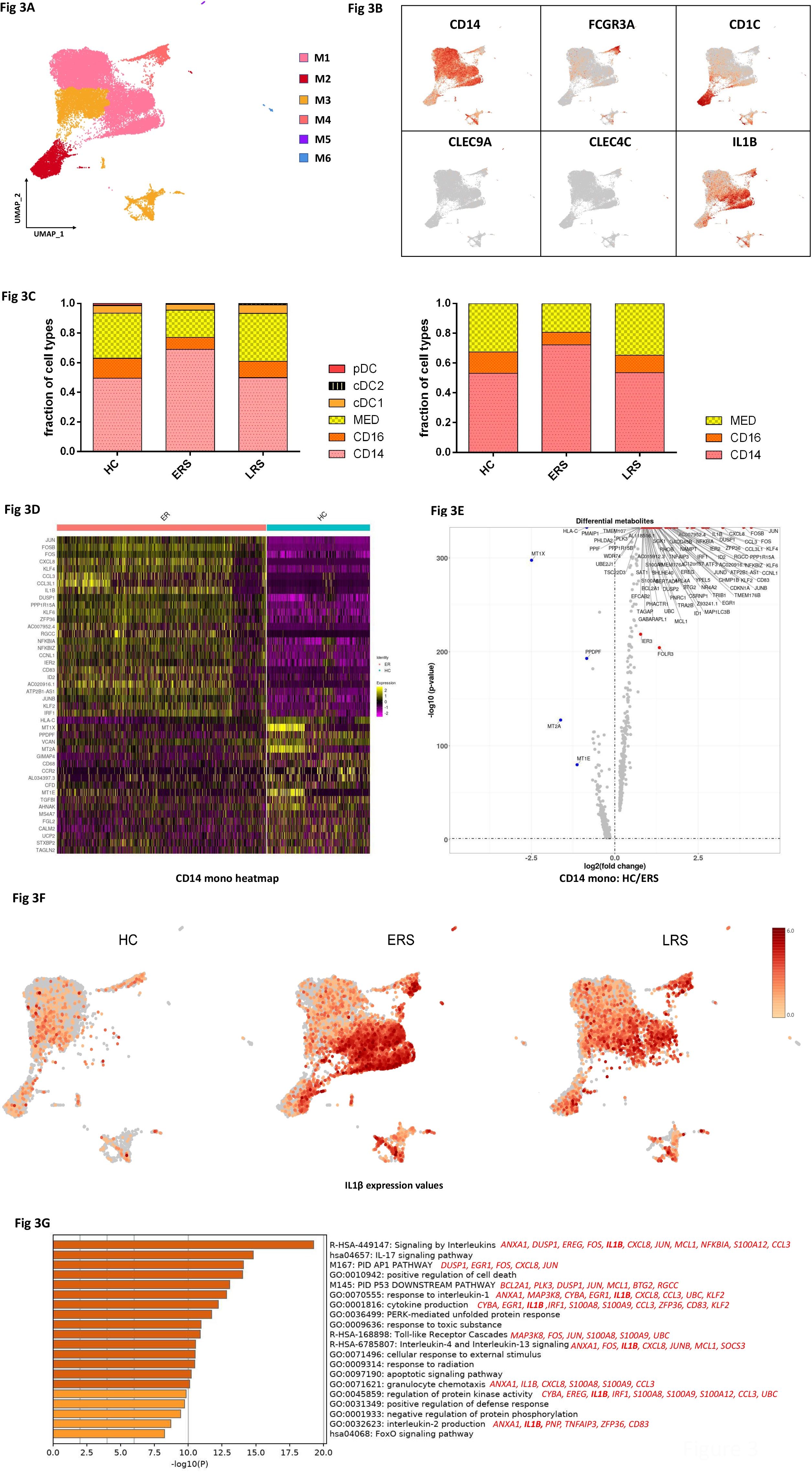
Myeloid cell subsets and their states in the blood of convalescent patients with COVID-19. A. Six clusters of myeloid cells were displayed according to marker gene expression levels. Uniform manifold approximation and projection (UMAP) presentation of the heterogeneous clusters of peripheral myeloid cells. B. The UAMP plot shows subtype-specific marker genes of myeloid cells, including *CD14, FCGR3A, CD1C, CLEC9A, CLEC4C*, and *IL-1β*. C.Bar chart of the relative frequencies of the six sub-clusters of myeloid cells and three sub-clusters of monocytes across the HCs and the ERS and LRS patients. D. The heatmaps show the top DEGs between COVID-19 patients and HCs in CD14^++^ monocytes. E. Volcano plot of fold change between COVID-19 patients and HCs in CD14^++^ monocytes. F The UAMP plot shows that IL-1β was highly expressed in the ERS patients vs. the LRS patients and HCs in myeloid cells. G. GO BP enrichment analysis of the DEGs of CD14^++^ monocytes upregulated in COVID-19 patients.

We found that COVID-19 patients had a greater abundance of CD14^++^ IL1β^+^ monocytes and IFN-activated monocytes than the HCs (Fig. 3D-F). Genes associated with CD14^++^ inflammatory monocytes (M1) had high expression levels of inflammatory genes such as *IL1*β, *JUN, FOS, JUNB*, and *KLF6*; chemokines, *CCL4, CXCR4*; and interferon-stimulated genes, *IFRD1, IRF1*, and *IFI6*. In contrast, anti-inflammatory genes associated with CD14^++^ monocytes (M1) were downregulated in COVID-19 patients relative to that in the HCs (Fig. 3D, E). Notably, IL1β expression values in a UMAP with simultaneous contrast indicated that IL1β was upregulated in the ERS group and decreased in the LRS patients (Fig. 3F). This was also confirmed in the DC cluster of the ERS group compared to that of the HCs (Fig. S3A-B). Next, we took the average of the inflammatory genes for each myeloid cell scRNA-seq subset in the COVID-19 patients versus that in the HCs (Fig. S3C). These results demonstrated that cytokine activation drives the expansion of monocyte populations (especially CD14^++^ inflammatory monocytes) in COVID-19-infected patients. To explore the biological significance of the transcriptional changes in the M1 cluster, we performed GO analysis with DEGs (Fig. 3G). We observed enrichment of the pathways related to cytokine signaling and inflammation activation, which were driven by the upregulation of *IFITM3* and *IFI6* and *IL1*β, *JUN, FOS, JUNB*, and *KLF6* (Fig. 3G).

Collectively, these findings demonstrate that a dysregulated balance in the monocyte populations in ERS patients is manifested by substantially increased classical CD14^++^ monocytes. Our results suggest that the classical CD14^++^ monocytes increase in circulation to fuel inflammation during SARS-CoV-2-infection.

### Characterization of T and NK cell responses in the blood of recovered COVID-19 patients

T and NK cells play critical roles in viral clearance during respiratory infections ^[15, 16]^. Our clustering analysis sub-grouped T and NK lymphocytes into 10 subsets (Fig. 4A) based on canonical markers (Fig. 4B and Fig. S4A). NK cells highly expressed *NCAM1, KLRF1, KLRC1*, and *KLRD1*; then, we sub-divided the NK cells into CD56^+^CD16^-^NK cells (NK1), which expressed high levels of *CD56* and low levels of *CD16*, and C56^-^CD16^+^ NK cells (NK2), which expressed high levels of *CD16* and low levels of *CD56*. CD4^+^ T cells expressed *CD3E* and *CD4*; then, we sub-divided these cells into four clusters: naïve CD4^+^T cells (T1), which expressed high levels of *CCR7, LEF1*, and *TCF7*; central memory CD4^+^T cells (T2, CD4 Tcm), which expressed high levels of *CCR7*, but more *AQP3* and *CD69* compared to naïve CD4^+^T cells; effector memory CD4^+^T cells (T3, CD4 Tem), which expressed high levels of *CCR6, CXCR6, CCL5*, and *PRDM1*; and regulatory T cells (T4, Treg), which expressed *FOXP3*. CD8+ T cells expressed *CD8A* and *CD8B* and were sub-divided into three clusters: naïve CD8^+^ T cells (T5), which expressed high levels of *CCR7, LEF1*, and *TCF7*, similar to naïve CD4^+^T cells; effector memory CD8^+^T cells (T6, CD8 Tm), which expressed high levels of *GZMK*; and cytotoxic CD8^+^ lymphocytes (CD8^+^ CTL) (T7), which expressed high levels of *GZMB, GNLY*, and *PRF1*. Proliferating T cells (T8, Pro-T) were *TYMS*^*+*^ *MKI67*^*+*^ cells.

**Figure 4.**
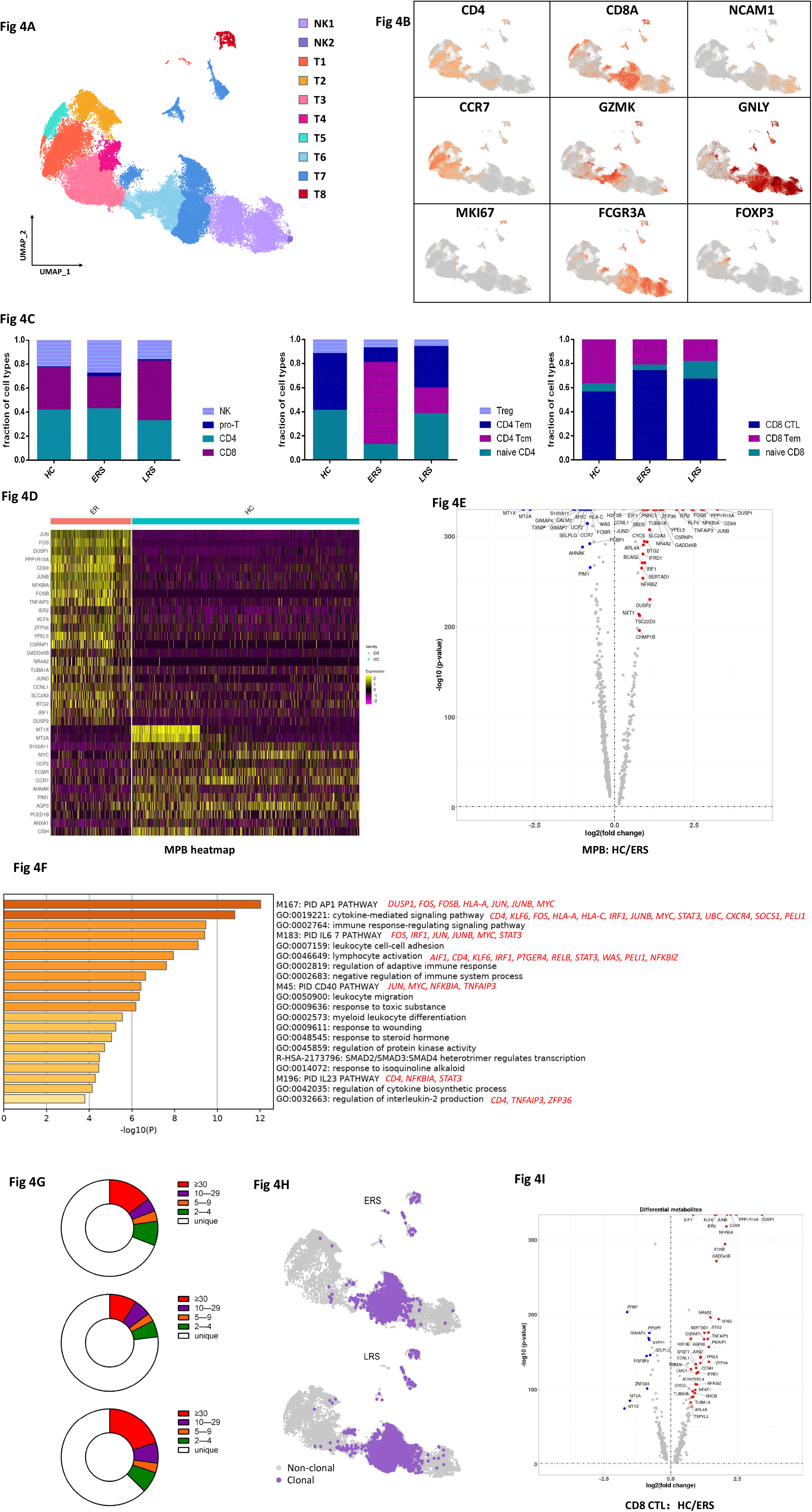
Characterization of T and NK cell responses in the blood of recovered COVID-19 patients. A. Ten sub-clusters of NKT lymphocytes were identified. The UMAP plot shows the clustering of T and NK cells. B. UAMP plot showing subtype-specific marker genes of NKT cells including *CD4, CD8A, NCAM1, CCR7, GZMK, GNLY, MKI67, FCGR3A*, and *IL-1*β. C. The bar plot shows the percentages of four sub-clusters of NKT cells, four sub-clusters of CD4^+^ T cells, and three sub-clusters of CD4^+^ T cells among the HCs and the ERS and LRS patients. D. Heatmap of CD4^+^ T cells showing the DEGs between the COVID-19 patients and HCs. E. The volcano plot shows the DEGs of CD8^+^ T cells between the COVID-19 patients and HCs. F. GO BP enrichment analysis of the DEGs of CD4^+^ T cells upregulated in the COVID-19 patients. G. The pie plot shows the TCR clone differences across the HCs and the ERS and LRS patients. H. UAMP shows expanded TCR clones (n≥2) in the ERS and LRS patients. I. The volcano plot shows the DEGs of CD8^+^ CTLs between the COVID-19 ERS group and HCs.

The composition of the T and NK cell subsets differed significantly among the HCs and COVID-19 patients (Fig. 4C). The ratio of CD8^+^T cells decreased in the ERS COVID-19 patients, whereas the ratio of NK cells was higher than that in the HCs. The ratio of CD4^+^ T cells was stable, but the composition of the CD4^+^ T cell subset differed significantly between the HCs and COVID-19 patients. Among CD4^+^ T cells, central memory CD4^+^ T cells were significantly higher, whereas the ratio of naïve CD4^+^ T cells was lower than thats in the HCs. Notably, genes associated with CD4^+^ T cells had relatively high expression levels of inflammation-related genes and were significantly upregulated in the COVID-19 patients (Fig. 4D). CD4^+^ T cells had high expression levels of inflammatory genes, includin*g FOS, JUN, KLF6*, and *S100A8* in patients in the ERS of COVID-19. (Fig. 4E). In contrast, anti-inflammatory genes associated with CD4^+^ T cells were downregulated in COVID-19 patients relative to that in the HCs (Fig. 4D, E). This suggested that CD4^+^ T cells were the main participants in the virus infection. Comparison of the DEGs in the CD4^+^ T cells revealed the enrichment of genes participating in the cytokine pathway and inflammation activation, including *IFITM3* and *IFI6* and *IL1B, JUN, FOS, JUNB*, and *KLF6* (Fig. 4F). Further studies are needed to elucidate the IFN pathways involved in COVID-19 pathogenesis.

TCR-seq analysis showed that T cell expansion was obviously decreased in the ERS group than in the HC group (Fig. 4G). Moreover, naïve or central memory T cells showed little clonal expansion, while effector memory T cells, terminal effector CD8^+^ T cells (CTLs), and proliferating T cells showed higher expansion levels (Fig. 4H). In addition, the most highly expanded (maximum) clone in the ERS group was TRAV8-6-TRAJ45:TRAV7-8-TRBJ2-1 (Fig. S5D). The decreased ratio of CD8^+^ T cells in COVID-19 patients may implicate the role of CD8^+^ T cells in virus clearance (Fig. 4C). Moreover, the CD8^+^ CTL with expanded clones also exhibited overactivated inflammation and antiviral activity compared to those in HCs (Fig. 4I and Fig. S4B). Together, these findings show that clonally expanded CD8^+^ T cells in the peripheral blood of COVID-19 patients help control the virus. We also performed DEG analysis, via unsupervised clustering analysis, and found an overactivated inflammatory state in pro-T cells (Fig. S4C). Next, we took the average of inflammatory genes for each NKT cell subset scRNA-seq subset in the COVID-19 patients versus normal RNA-seq data (Fig. S4D).

### Characterization of single-cell B cells in COVID-19 patients

By projecting the gene expression data of B cells using diffusion maps, we identified four B cell clusters using scRNA-seq: naïve B cells (B1) expressing *CD19, CD20 (MS4A1), IGHD, IGHM, IL4R*, and *TCL1A*; memory B cells (B2) expressing *CD27, CD38*, and *IGHG*; immature B cells (B3) only expressing *CD19* and *CD20 (MS4A1)*; and plasma cells (B4) expressing high levels of *XBP1* and *MZB1* (Figs. 5A-B and Fig. S5A).

**Figure 5.**
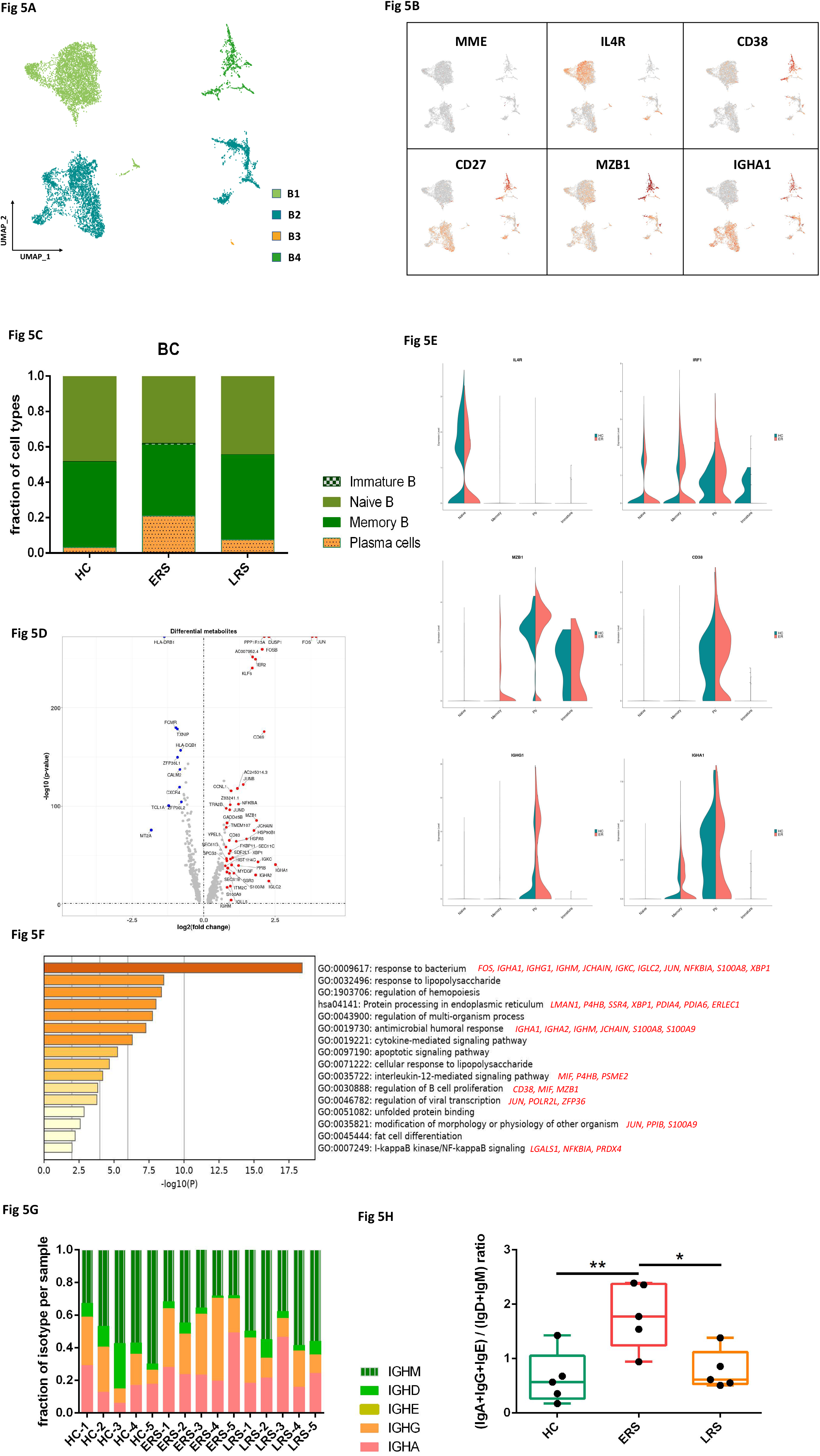
Characterization of single-cell B cells in COVID-19 patients. A. Four clusters of B cells were identified. The UMAP plot shows the clustering of B cells. B. UAMP plot showing subtype-specific marker genes of B cells, including *MME, IL4R, CD38, CD27, MZB1*, and *IGHA1*. C. The bar plot shows the percentages of B clusters across the HCs and the ERS and LRS patients. D. The volcano plot shows the DEGs of MPB cells between the COVID-19 patients and HCs. E. The violin plot shows that *IL4R, MZB1, IGHG1*, and *IGHA1* were highly expressed in COVID-19 patients vs. the HCs in the B cell sub-clusters. F. GO BP enrichment analysis of the DEGs of B cells between the COVID-19 patients vs. the HCs. G. The bar plot shows the relative contributions of each cluster by individual sample. H. The bar plot shows the ratio of (IgA+IgG+IgE) to (IgD+IgM) among the HCs and the ERS and LRS patients.

In comparison with that in the HCs, the percentage of plasma cells increased significantly in COVID-19 patients, whereas naïve B cells decreased significantly in the COVID-19 patients (Fig. 5C). Memory B cells and plasma cells (MPB) might play an important role in the control of viral infection and the development of adoptive immunity as they synergistically work and induce specific antibodies. Moreover, compared to that in the HCs, B cell activation-related genes, including *S100A8, IGLL5, SSR3, IGHA1, XBP1*, and *MZB1* were primarily expressed in the MPB of the ERS group (Fig. 5D). We also found similar results in the plasma cells, the antibody-secreting cells (ASC) (Fig. S5B-C), suggesting a key role for ASC in viral control. Next, we took the average of the inflammatory genes for each B cell subset of the COVID-19 patients versus the normal RNA-seq data (Fig. 5E). The difference in the genes between the ERS and HCs indicated enhanced B cell reaction and antibody secretion in COVID-19 patients. GO analysis revealed that *IGHA1, XBP1, MZB1, JUN, POLR2L*, and *ZFP36* were over-presented in MPBs, which suggests enhanced B cell proliferation and viral transcription in COVID-19 patients (Fig. 5F). Single-cell BCR-seq analysis indicated that the IgA isotype was over-represented in COVID-19 patients compared to that in the HC (Fig. 5G). This corresponded with an increase in the levels of serum IgA, which was also pronounced in other coronavirus infections. Moreover, the ratio of (IgA+IgG+IgE) to (IgD+IgM) increased significantly in the ERS patients and showed a downward trend with recovery time (Fig. 5H).

### Expanded BCR clones and biased usage of VDJ genes observed in COVID-19 patients

Using sc-BCR-seq to assess the status of clonal expansions in the blood of patients, we found that IL4R^+^ naïve B cells showed little clonal expansion, whereas CD27^+^CD38^+^ memory B cells showed the highest expansion levels among diverse B cell subsets (Fig. 6A). At the individual level, we found that COVID-19 patients had significantly expanded clones compared to that in the HCs, supporting the assumption that B cells had experienced unique clonal VDJ rearrangements under SARS-CoV-2 infection. We also found that a higher B cell clonality consistently remained in the ERS compared with that in the LRS patients (Fig. 6B). Moreover, quantification of the most highly expanded (maximum) clone for each subject showed that the ratios of the maximum clones were higher in the ERS group than in the HCs (Fig. 6C). To understand the functional status of expanded cloned B cells, we performed DEG analysis between the cloned memory B cells and the other B cells.

**Figure 6.**
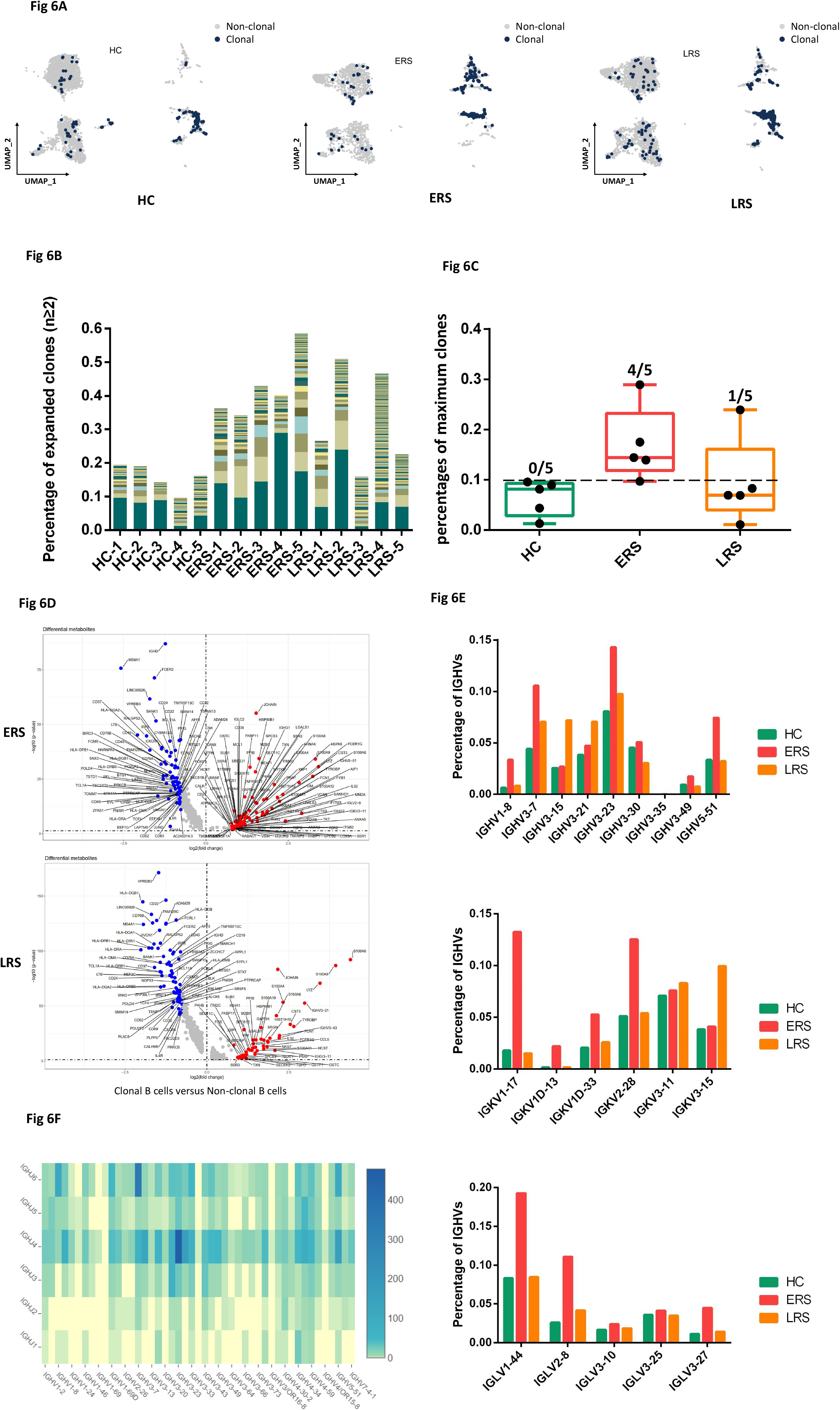
Expanded BCR clones and biased usage of VDJ genes observed in the COVID-19 patients. A. The UMAP plot shows the B cell expansion status in the HCs and the ERS and LRS COVID-19 patients. B. The bar plots show the clonal expansion status of B cells in peripheral blood from each individual sample. C. The bar plots show the percentages of maximum clones of B cells in the peripheral blood of the HCs and the ERS and LRS COVID-19 patients. D. The volcano plot shows the DEGs of expanded vs. non-expanded B cells in ERS and LRS patients. E. The bar plots show specific IGHV, IGKV, IGLV usage in the HCs and the ERS and LRS COVID-19 patients. F. Heat map showing IGH rearrangements in peripheral blood samples from ERS group.

Our results revealed increased expression of B cell genes, including *CD27, SSR4, IGHG1, MZB1*, and *XBP1*, which further supports the superior effector functions of the expanded cloned B cells (Fig. 6D). Moreover, the differential genes for expanded B cells significantly subsided over time and reduced in LRS patients (Fig. 6D).

To study the unique changes and preference genes of BCR in COVID-19 patients, we compared the usage of VDJ genes in COVID-19 patients with that in the HCs. We identified an over-representation of the IGHV3 family, especially the IGHV3-7, IGHV3-15, IGHV3-21, IGHV3-23, and IGHV3-30 in COVID-19 patients compared to that in the HCs (Fig. 6E). The preferred IGLVs were IGKV1-17, IGKV2-28, and IGKV3-15, whereas the preferred IGKVs were IGLV1-44, IGLV2-8, and IGLV3-27 (Fig. 6E). Moreover, the top two pairing frequencies in ERS patients were IGHV3-23-IGHJ4 and IGHV3-7-IGHJ6 (Fig. 6F). These cells showed IGH subunit pairing with the IGL/V subunit encoded by IGLV1-44-IGLJ3 and IGKV1-17-IGKJ1, respectively, which indicated expanded states associated with SARS-CoV-2 specificity. Individually, ERS-4 and ERS-5 had the maximum clones, referring to IGHV3-23-IGHJ4 (Fig. S5E) and IGHV3-7-IGHJ6 (Fig. S5F), respectively.

In summary, an increase in clonality in COVID-19, which was dominated by the IgA and IgM isotypes, together with a skewed use of the IGHV gene, suggested the contribution of SARS-CoV-2 to pathogenesis. Notably, the biased usage of dominated IGV genes, especially the IGHV3-23 and IGHV3-7 in COVID-19 patients, provides a framework for the rational design of SARS-CoV-2 vaccines.

### Cell-to-cell communication among immune cells in COVID-19 patients

An established computational approach ^[17]^ was used to predict cell-to-cell interactions that may contribute to the distinct functional state of T cells, B cells, monocytes, and dendritic cells (DCs) in ERS and LRS (Fig. 7A, B). In ERS COVID-19 patients, we found adaptive signals involved in monocyte activation, proliferation, and inflammatory signaling (Fig. 7A, B). T cells expressed genes encoding ligands of TNFSF8, LTA, IFNG, IL17A, CCR5, and LTB to TNFRSF8, TNFRSF1A/TNFRSF14, IFNGR1, IL-17RA, CCR1, and LTBR, which were expressed on monocytes and could contribute to the pro-inflammatory status. Other T cell-monocyte interactions involved the expression of CSF2 and CSF1. T cells might activate monocytes through the expression of CSF2 and CSF1, which bind to CSFRs (CSFR2/1) and contribute to inflammatory storm. A cluster of CD14^+^ monocytes exclusively expressed IL1β, which was predicted to bind to IL1RAP expressed by T cells. T cell-monocyte interaction may enhance immune response and be exclusive to COVID-19 patients. (Fig. 7A, B). Furthermore, we found that monocytes highly expressed the poliovirus receptor, which serves as a cellular receptor for poliovirus in the first step of poliovirus replication and induction of the NF-kappa B signaling pathway. From the B cell-monocyte and B cell-T cell interactions, we found that B cells could secrete a large number of IL-6, LTA, and LTB, which are combined with IL-6R, LTAR, and LTBR expressed in monocytes, and a large amount of IL-6 was applied to T cells to promote the secretion of IFN-γ, IL-1β, and other inflammatory cytokines and chemokines. Thus, a cascade signature of inflammatory monocytes with high expression of IL-6 and their progeny were formed in the peak incidence of ERS COVID-19 patients (Fig. 7C). These activated immune cells may enter the circulation in the lung and other organs in large numbers and play an immune-damaging role. In LRS COVID-19 patients, DC ligands were predicted to interact with B and T cell receptors involved in cell proliferation and the production of antibodies. We discovered that the peripheral blood of LRS patients contains a diversity of antibodies; we found that IL18-IL18RAP, TNFSF13-TNFRSF13B, TNFSF13-TNFRSF17, TNFSF13B-TNFRSF17, TNFSF13B-TNFRSF13B, and TNFSF13B-TNFRSF13C were highly expressed in our analysis of DC-B cell interaction (Fig. 7D). Thus, we speculate that DCs produce IL-18, TNFSF13, and TNFSF13B to promote the proliferation of B cells and then secrete many antibodies into the blood in ERS. From the DC-T and T cell-B cell interactions, we discovered that DCs produce not only IL-18 but also IL-7 to promote the proliferation of T cells; moreover, T cells produce IL-2 (to promote the proliferation of B cells) and antibodies (Fig. 7D). Thus, cell-to-cell interactions help us to understand why COVID-19 patients manifested high rates of monocytes and low rates of lymphocytes and why the proportion of lymphocytes gradually increased in the peripheral blood of recovering patients.

**Figure 7.**
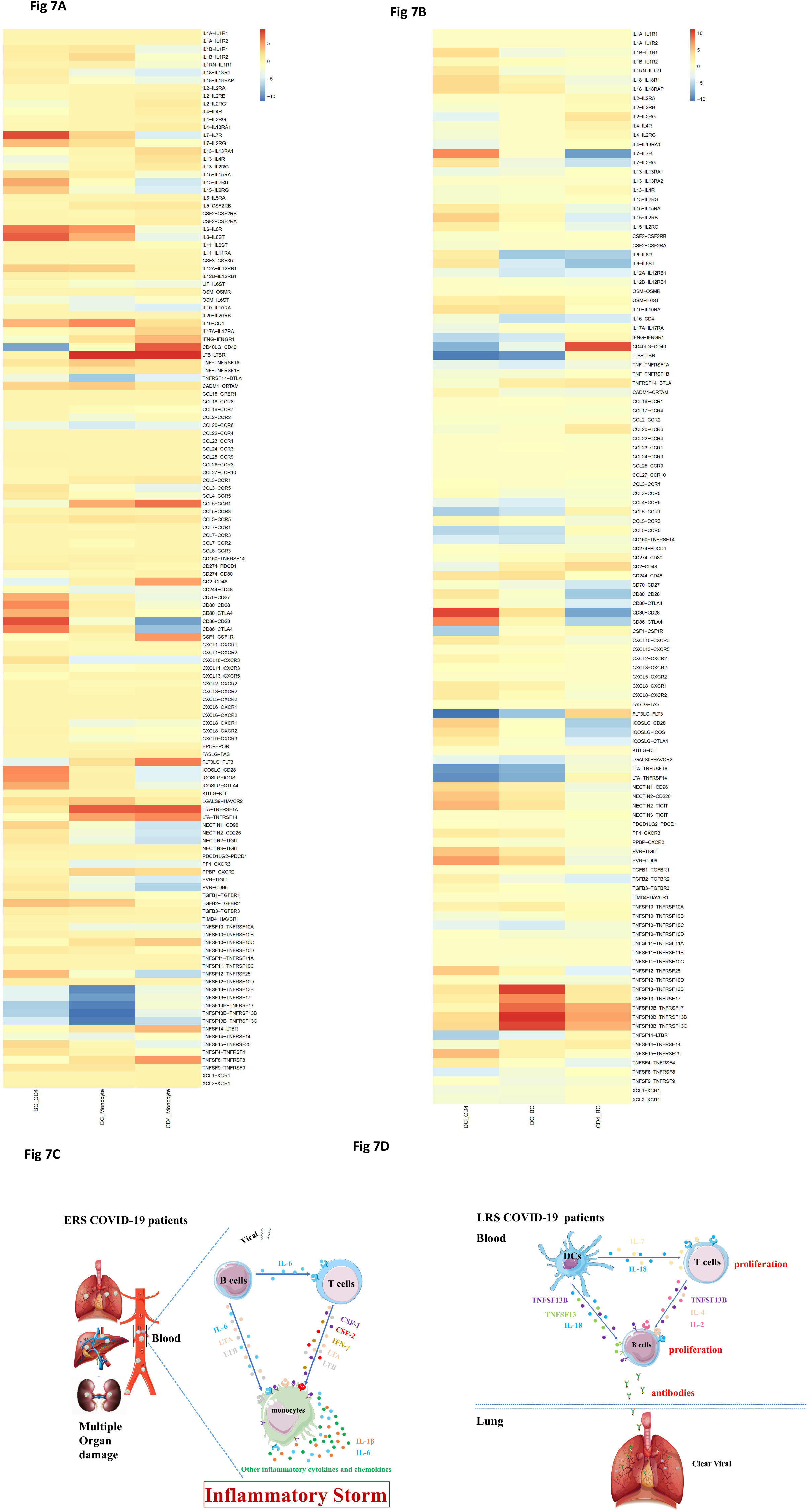
Cell-to-cell communication among immune cells in the COVID-19 patients. A. T cell-monocyte interactions, B cell-monocyte interactions, B cell-T cell interactions, and monocyte-T cell interactions in the ERS COVID-19 patients. B. DC-T cell interactions, DC-B cell interactions, and T cell-B cell interactions in the LRS COVID-19 patients. C-D. Schematics illustrating the key innate and adaptive immune cell functional alterations and main differences in cell-cell communications in the ERS and LRS COVID-19 patients.

## Discussion

The clinical presentation of COVID-19 varies from asymptomatic to severe ARDS. This has been similarly observed in severe acute respiratory syndrome coronavirus (SARS-CoV), Middle East respiratory syndrome coronavirus (MERS-CoV), and influenza infections ^[12, 14]^. In viral infection, it is generally accepted that host immune responses determine both protection against viral infections and the pathogenesis of respiratory injury ^[18, 19]^. A coordinated response in innate and adaptive immune cells working in concert may lead to the rapid control of the virus, whereas a failed immune response might lead to viral spreading, cytokine storm, and a high mortality rate ^[20]^. Despite belonging to same group of viruses, recent studies have highlighted differences between COVID-19, SARS, and MERS, such as the speed of transmission, treatment scheme, and mortality rate. Moreover, this difference may also exist in the key immune players and the underlying molecular mechanisms related to these diseases. The lack of knowledge regarding the immune impact of COVID-19 has now become a critical issue in view of its rapid spread and the shortage of specific therapy ^[21]^. Using single-cell sequencing, we profiled the complexity of immune populations in the blood and analyzed 70,858 cells from 10 patients. We identified a hyper-inflammatory response in ERS patients, which may explain why some patients fell sick after being discharged, and suggest that the current criteria for hospital discharge should be re-evaluated. In addition, we identified unique signatures of myeloid, NKT, and B cells and pinpointed the changes in the epitopes of TCR and BCR. Our findings helped elucidate the antiviral immune mechanisms and revealed promising opportunities for developing immunotherapies using vaccines and neutralizing antibodies.

Inflammation is a vital part of the immune system’s response to COVID-19 invasion; previous and latest studies have reported significantly higher levels of inflammatory cytokines associated with disease severity in SARS, MERS, and COVID-19 patients^[22, 23]^. Among the various inflammatory cells, monocytes and their subsets (including classical, intermediate, and non-classical monocytes) may play a critical role because they are known to fuel inflammation ^[24-27]^. In our study, compared with the HCs, ERS patients demonstrated a significantly higher ratio of monocytes, and these cells expressed higher levels of inflammatory genes. Intriguingly, the ratio of classical CD14^+^ monocytes was high in ERS but remained normal in LRS. Furthermore, CD14^+^IL1β monocytes, which were absent in HCs, could be observed in ERS, and they declined in number in LRS. Notably, our cell-to-cell interaction analysis indicated that IL1β, CSF1, IL6, and CSF2 may be associated with cytokine storm. Collectively, our data provide important insights into the role of monocytes in the immunopathogenesis of COVID-19.

The adaptive immune system harbors the ability to recognize and remember specific pathogens through antibody and T cell responses ^[28]^. Inducing adaptive immunity is the aim of vaccination ^[29]^. Previous SARS studies have identified binding and neutralizing antibodies elicited by SARS-CoV infection. Their therapeutic effect is unclear ^[30]^, although robust antibody responses could be induced ^[31]^. In COVID-19 infection, although several lines of evidence have consistently indicated a decline in lymphocyte counts, the distinct immune characteristics at single-cell resolution are unclear. Our scRNA-seq analysis showed that, compared with the HCs, ERS patients have a lower ratio of T and NK cells, and these patients’ T cells express higher levels of inflammatory genes, such as *JUN, FOS, JUNB*, and *KLF6*. In addition, high-throughput TCR sequencing identified expanded T cell clones in ERS patients. In LRS patients, the immunophenotype was different. In particular, LRS patients would have an increase in T and NK cells, with a lower expression of inflammatory genes. We also performed a detailed analysis of B cells in patients and identified a higher population of plasma cells than that in the HCs. We found that BCR contained highly expanded clones, indicating their SARS-CoV-2 specificity. Importantly, we found several loci unique to COVID-19 infection. The strongest pairing frequencies, IGHV3-23-IGHJ4, indicated a monoclonal state associated with SARS-CoV-2 specificity. Notably, numerous studies have reported biased usage of VDJ genes related to virus-specific antibodies. For example, IGHV3-30 and IGKV3-11 have been involved in encoding primary antibodies to neutralize human cytomegalovirus^[32, 33]^. In addition, IGHV3-30 and IGHV3-21 have been utilized to isolate influenza virus antibodies and used for the production of virus vaccines ^[34, 35]^. Moreover, a recent study demonstrated that antibodies combining the IGHV3-15/IGLV1-40 segments had superior neutralizing activities against the Zaire Ebola virus^[36]^. In addition, we observed lower expression of inflammatory genes in ERS patients than in the HCs. We envision that our results will provide direction for the development of vaccines and antibodies for COVID-19 patients.

Interaction between immune cells may help expedite or defer recovery from COVID-19 infection. Our cell-to-cell prediction analysis utilizing scRNA-seq data indicated that, in ERS patients, B cell-derived IL-6, T cell-derived CSF1 (M-CSF), and CSF2 (GM-CSF) may promote monocyte proliferation and activation. As a result, monocytes may produce a larger number of inflammatory mediators, including IL-1β and IL-6, contributing to inflammatory storm. In LRS patients, both DCs-derived TNFSF13 and IL-18 and T cell-derived IL-2, IL-4 may promote B cell survival, proliferation, and differentiation. Consequently, B cells produce numerous SARS-COV-2-specific antibodies to clear viruses.

In conclusion, our study provides the first immune atlas of patients who have recovered from COVID-19 and identifies adaptive immune dysregulation after discharge. The clonal expansion of both T and B cells indicates that the immune system has gradually recovered; however, the sustained hyper-inflammatory response for more than 7 days after discharge suggests the need for medical observation after patients are discharged from hospital. Longitudinal studies of recovered patients in a larger cohort might help to understand the consequences of the disease. The novel BCRs identified in our study may advance our understanding of B cell mechanisms and have potential clinical utility in COVID-19 immunotherapies.

## Methods

### Software and Algorithms

|  |  |
| --- | --- |
| Prism 8 | https://www.graphpad.com/scientificsoftware/prism/ |
| Cell Ranger 3.1.0 | https://support.10xgenomics.com |
| Loupe Browser 4.0.0 | https://support.10xgenomics.com |
| Loupe V(D)J Browser 3.0.0 | https://support.10xgenomics.com |
| R 3.6.1 | https://www.r-project.org/ |
| Seurat V3<sup>[37]</sup> | https://satijalab.org/ |
| Monocle3<sup>[38]</sup> | https://cole-trapnell-lab.github.io/monocle3 |
| GO analysis<sup>[39]</sup> | https://www.metascape.org |

### Patients

10 COVID-19 patients diagnosed with by real-time fluorescent RT-PCR were collected in the Wuhan Hankou Hospital China. Patients were divided into early-recovery stage (ERS) group and late-recovery stage (LRS) group according to the days from first negative nucleic acid transfer date to blood sampling date. We defined the RES group of five cases as the date of nucleic acid turning negative to blood sampling is less than seven days and LRS group of five cases as is more than fourteen days. The 10 patients consisted of five males and five females and ranged from ages 40 to 70 years old, with a median of 50 years old. The demographic characteristics of these patients are provided in Fig. S1. A written informed consent was regularly obtained from all patients. The study was approved by the Ethics Committee of Wuhan Hankou Hospital, China.

### Quantitative reverse transcription polymerase chain reaction

The throat swab, sputum from the upper respiratory tract and blood were collected from patients at various time-points after hospitalization. Sample collection, processing, and laboratory testing complied with WHO guidance. Viral RNA was extracted from samples using the QIAamp RNA Viral Kit (Qiagen, Heiden, Germany) according to the manufacturer’s instructions. SARS-CoV-2-infected patients were confirmed by use of qRT-PCR kit (TaKaRa, Dalian, China) as recommended by China CDC.

### Single-cell collection and scRNA-seq

The peripheral blood mononuclear cell (PBMCs) were isolated from heparinized venous blood of patients or healthy donors using a Ficoll-Hypaque density solution according to standard density gradient centrifugation methods. For each sample, the cell viability exceeded 80%.

The single-cell suspensions of scRNA-seq samples were converted to barcoded scRNA-seq libraries using the Chromium Single Cell 5′ Library, Gel Bead and Multiplex Kit, and Chip Kit (10X Genomics). The Chromium Single Cell 5′ v2 Reagent (10X Genomics, 120237) kit was used to prepare single-cell RNA libraries according to the manufacturer’s instructions. The FastQC software was used for quality check. The CellRanger software (version 3.1.0) was used for initial processing of the sequencing data.

### ScRNA-seq data alignment and sample aggregating

We de-multiple and barcode the sample by using The Cell Ranger Software Suite (Version 3.1.0) and with command cellranger count. After getting each sample gene counts, and aggregate them together. Finally, gene-barcode matrix of all ten patients and five HCs was integrated with Seurat v3 and monocle3. Following criteria were then applied to each cell, i.e., gene number between 200 and 7000. After filtering, a total of 128096 cells (13092/10035/13624/8329/12158 cells for HCs; 5163/7685/7171/10058/6581 cells for ERS; 3242/7895/7487/7164/8412 cells for LRS) were left for following analysis.

### Dimensionality reduction and clustering

We handle the data with Log normalize before cluster and reduction, scale data with top 5000 most variable genes. Clustering and dimensionality method mainly used in monocle3 package. The genes used in PCA analysis have eliminated mitochondria (MT), and ribosomes (RPL and RPS) genes including MT-ND3, MT-ATP8, RPS15A, RPS28, RPS21, RPS27, RPS29, RPL36, RPL34, RPL37, RPL38, RPL39, RPL26 and et.al. with 50 principal components, and then aligned together, followed by UMAP and t-SNE are both used after the results of the aligned, parameters using the default parameters inside Monocle3. The leiden method on the UMAP dimension is used.

### Differential analysis for clusters

Seurat v3 and Monocle 3 was used to perform differential analysis. For each cluster, differentially-expressed genes (DEGs) were generated relative to all of the other cells.

### Gene functional annotation

For DEGs, Gene ontology (GO), KEGG pathway analyses were performed using Metascape webtool (www.metascape.org).

### TCR and BCR V(D)J sequencing and analysis

Full-length TCR/BCR V(D)J segments were enriched from amplified cDNA from 5’ libraries via PCR amplification using a Chromium Single-Cell V(D)J Enrichment kit according to the manufacturer’s protocol (10x Genomics). The TCR/BCR sequences for each single T/B cell were assembled by Cell Ranger vdj pipeline (v3.1.0), leading to the identification of CDR3 sequence and the rearranged TCR/BCR gene. Analysis was performed using Loupe V(D)J Browser v.2.0.1 (10x Genomics). In brief, a TCR/BCR diversity metric, containing clonotype frequency and barcode information, was obtained. Using barcode information, T/B cells with prevalent TCR/BCR clonotypes were projected on a t-SNE plot.

### Cell-cell interaction analysis

The cell-cell interaction analysis was based on the expression of immune-related receptors and ligands. The gene list contained 135 pairs of well-annotated receptors and ligands, including cytokines, chemokines and co-stimulators. We estimated the potential interaction between two cell types mediated by a specific ligand-receptor pair by the product of the average expression levels of the ligand in one cell type and the corresponding receptor in the other cell type.

## Data Availability

https://bigd.big.ac.cn

## Acknowledgements

This study was supported by National Natural Science Foundation of China (Grant No. 81722034, 81670015, 81830054, 81530028, 91953122, 31871326, 81721003, 81988101), the Ministry of Science and Technology Key Program (Grant No. 2018ZX09101002, 2017ZX100203205), Shanghai Pujiang Program (Grant No. 2019PJD059), and Guangdong Natural Science Funds for Distinguished Young Scholar (Grant No. 2016A030306006, 2020B1515020057).

## Figure legends

**Supplementary figure 1**. the detail information of patients (1A) and healthy controls (1B).

**Supplementary figure 2**. tSNE plot showing the myeloid cells (red), NK&T cells (blue), B cells (green) and other clusters (grey) of PBMCs of each research object.

**Supplementary figure 3**. Analysis myeloid cells subsets landscape in COVID-19 patients. The heatmap of DCs showing the DEGs between COVID-19 patients and HCs (3A). The volcano plot shows the DEGs of DCs between COVID-19 patients and HCs (3B). The violin plot shows the *JUNB, FOS, S100A8, ISG15, IRF1, IFI6, CXCR4, IL1* β, *CD83* were highly expressed in COVID-19 patients vs HCs in myeloid cells (3C).

**Supplementary figure 4**. Analysis NK&T subsets landscape in COVID-19 patients. The UAMP plot showing subtype-specific marker genes of NK&T cells including *KLRF1, KLRC1, PRF1, CCL5, TCF7, LEF1, CD69, CD27, CTLA4, CCR6, GZMB and TYMS* (4A). GO BP enrichment analysis of the DEGs of CD8^+^ CTLs between the COVID-19 patients vs. the HCs. (4B). The volcano plot shows the DEGs of ERS COVID-19 patients vs. HCs in proliferating T cells (4C). The violin plot shows the *IRF1, STAT3, MKI67, ISG15, IFI6 and IFNG* were highly expressed in ERS COVID-19 patients vs HCs in T & NK sub-clusters (4.4).

**Supplementary figure 5**. Analysis B cell subsets landscape in COVID-19 patients. The UAMP plot showing subtype-specific marker genes of B cells including TCL1A, IGHM, IGHG1, XBP1, IGHD and MS4A1 (5A). The heatmap of ASCs showing the DEGs between COVID-19 patients and HCs (5B). The volcano plot shows the DEGs of COVID-19 patients vs. HCs in plasma B cells (5C). Heat map showing TRA and TRB rearrangements in peripheral blood samples from ERS group (5D). Heat map showing IGH rearrangements in peripheral blood samples from ERS-4 sample (5E). Heat map showing IGH rearrangements in peripheral blood samples from ERS-5 sample (5F).

